# Machine Learning Analysis of Chest CT Scan Images as a Complementary Digital Test of Coronavirus (COVID-19) Patients

**DOI:** 10.1101/2020.04.13.20063479

**Authors:** Dhurgham Al-Karawi, Shakir Al-Zaidi, Nisreen Polus, Sabah Jassim

**Affiliations:** Medical Analytica Ltd, 26A Castle Park Industrial Estate, CH6 5XA, Flint, UK; School of Computing, University of Buckingham, Buckingham, MK18 1EG, England, UK

**Keywords:** COVID-19, CT Scan, Machine learning, Fast Fourier Transform, Gabor Filter

## Abstract

This paper reports on the development and performance of machine learning schemes for the analysis of Chest CT Scan images of Coronavirus COVID-19 patients and demonstrates significant success in efficiently and automatically testing for COVID-19 infection. In particular, an innovative frequency domain algorithm, to be called FFT-Gabor scheme, will be shown to predict in almost real-time the state of the patient with an average accuracy of 95.37%, sensitivity 95.99% and specificity 94.76%. The FFT-Gabor scheme is adequately informative in that clinicians can visually examine the FFT-Gabor feature to support their final diagnostic.

**Key Strengths:** The proposed FFT-Gabor scheme is an automatic machine learning scheme that works in real time and achieves significantly high accuracy with very low false negative, and can provide supporting evidences of the predicted decision by visually displaying the final features upon which decision is made. This scheme will be most beneficial when used in addition to the RT-PCR swab test of non-symptomatic cases.

## 1. Introduction

Over the past few months, many countries in different continents have been feeling the devastating effects of the novel Coronavirus “SARS-CoV-2” with the number of confirmed cases surpassing 1.6 million world-wide with nearly 100,000 deaths [1]. Currently, there is no proven drug treatment or vaccine available for this virus. As of April 2020, WHO has identified more than 60 vaccine candidates being investigated against the SARS-CoV-2 virus ([2], [3]), but only 2 are undergoing clinical trials. As for Diagnostic testing for the SARS-CoV-2, almost all diagnostic testing is done using RT-PCR. Due to the asymptomatic and pre-symptomatic transmission nature of this virus, a growing demand for diagnostic testing is a serious challenge even for developed countries. Various serological tests have been developed and approved in some Asian, European countries and America [4]. The reliability of both types of tests and in particular in asymptomatic and pre-symptomatic patients has been brought to question due the number of reported false negatives. Therefore, radiology units in hospitals would have been naturally involved in assessing level of infection using their various scanning images (e.g. CT and Ultrasound). Ai, et al, manually examined Chest CT scan images of 1014 patients and demonstrated that such a scan provides a reliable tool for the diagnosis of COVID-19 infection [5]. More interestingly, they found that a number of patients had positive Chest CT findings despite having an initial negative PCR test thus confirming the diagnosis of COVID-19.

In [6], Zhong J et al, independently created and made public another COVID-19 CT scan dataset, to foster the use of AI methods for screening and testing COVID-19 patients. They proposed a Deep learning scheme that used the ChestX-ray14 dataset [7], to pretrain the DenseNet [8], then used Transfer learning to train and test samples of their dataset. Their scheme achieved accuracy of 84.7%. Earlier, Xu X. et al, [9] developed another Deep Learning based scheme to screen for COVD-19 pneumonia in a multi-centre case study. This scheme adds more evidence to the viability of using AI and machine learning as a supplementary diagnostic method for frontline clinical doctors. They achieved an overall accuracy of 86.7% for three groups: COVID-19, Influenza-A viral pneumonia and healthy cases.

In this paper, a new approch based on texture analysis has been develped to distinguish between positive from negative cases using a dataset of COVID-19 Chest CT scan images. In section 2, we shall first describe the background to our approach and illustrate with examples the viability of our developed machine learning schemes. In section 3, we shall present and discus the results of our experimental tests on the dataset created and frequently updated by the authors of [6] to whom we are indebted for their dedication.

## 2. FFT-based Classification of COVID-19 CT Scan Images

A computer-based system for categorising chest CT scans could contribute to improve performance of decision support systems for the diagnosis of COVID-19. Such a computer-based system benefits from an interdisciplinary technology that combines image processing methods and experts’ knowledge for greatly, better accuracy of positive detection, hence greatly decreasing the false-negative rate and improving the true positive rate. Recent technology advances in computer vision and machine learning have led to the successful development of automated analysis software tools as an integral of diagnostic support systems. Motivated by recent work, conducted by the first and last authors, on classifying Utrasound Ovarian Tumours by machine learning texture analysis schemes ([10], [11], [12], [13]). The main innovative contribution of this paper is the Gabor filtering of the spectrum image of the Fast Fourier Transform [13] of COVID-19 CT scan images as the domain of texture analysis for effectively differentiating positive from negative cases. The rationale is that all detailed texture information of images is extractible from the frequencies around the centre of the spectrum.

Fourier transform is a well-established mathematical tool that represents data functions/signals in terms of waveforms of different frequencies, and when applied to image data its frequency ranges are analysed in the same way as a prism analyses natural light into a rainbow. Higher frequency ranges are associated with image texture features. Human observers, including trained radiologists, might find it difficult to detect informative features from FFT spectrum images. However, a wealth of research on use of machine learning for image analysis provide strong evidences that FFT indeed incapsulate rich characterising texture information that discriminating power when used for classifying tissue/organ abnormality. There are many different known procedures to analyse the texture content of the FFT spectrum. However, the only texture analysis tool that can effectively determine the nature of the lines out of the central region in the FFT spectrum is the Gabor filter [14], which is a band-pass spatial filter that determine specific frequency content in the image in chosen ranges of directions and scales in the region of analysis. Figure 1 illustrates the output of using the FFT-Gabor scheme as shown of filtering the Fourier spectrum image in 5 scales and 10 orientations. It is easy to visibly note the differences in the pattern displayed in each of the 50 small FFT-Gabor blocks between the positive and negative COVID-19 cases.

**Figure 1.**
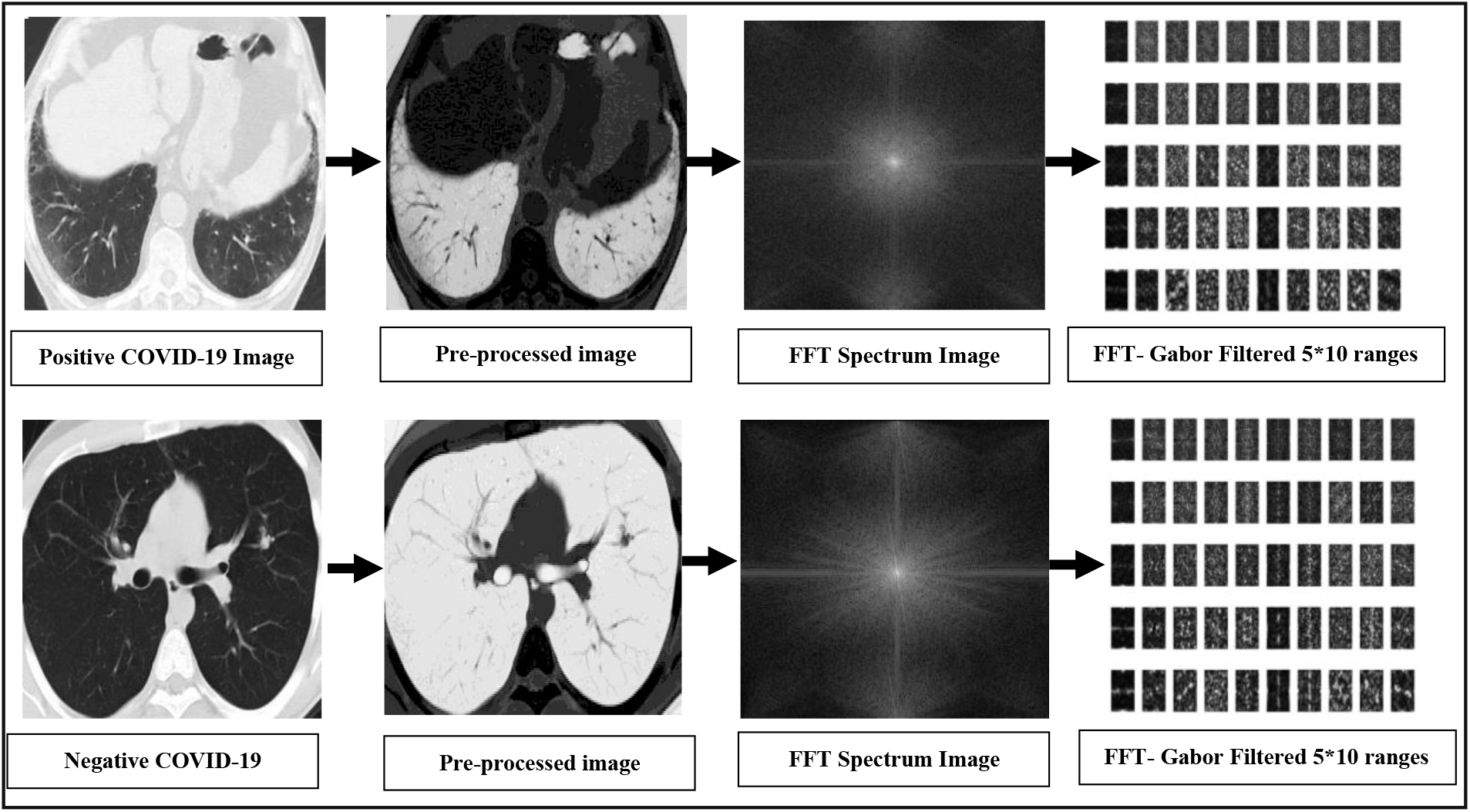
Examples of FFT-Gabor processing of positive and negative CT scan cases.

These observations were our strong motivation to investigate and design machine learning based schemes using extracted image texture features and test their performances on a sufficiently large dataset of CT scans in terms of distinguishing positive Covid-19 cases from negative ones. Automated FFT-based schemes works by (1) pre-process the image using adaptive winner filter for noise reduction and image quality improvement, followed by inversion, (2) obtain the FFT-spectrum, (3) select a texture type and extract the appropriate feature vector, and (4) train and test the performance of each scheme on a sufficiently large dataset of image, using the linear Support Vector Machine (SVM) classifier [13]. We choose not to use Deep Learning schemes that normally predict decision in a *black-box* manner, because we can provide convincing visible informative evidence to support clinical decisions.

## 3. Experimental Result and Discussion

In this study, chest CT-scan images of COVID-19 examined patients (275 positive and 195 negative) were downloaded from [6]. Since, the authors in [6] are frequently updating the image database by uploading batches of lung CT scans of newly examined patients, then we have decided to first publish the performance of our proposed schemes by training and testing the original 470 labelled samples using support vector machine (SVM) classifier, and each time there is a dataset update we select a randomly a 100 images from the new sets and test the performance of the existing classification model on the new 100 image akin to a prospective clinical testing. If the results did not change significantly then wait for the next set, otherwise we retrain using a mix of old and new images.

The left part of Figure 2, below, illustrates the experimental protocol used to test the performance of different schemes when we used the initial set of 470 downloaded images, whereas the right side illustrates a prospective test-like experiment where we using 100 images downloaded from the same website after second round of image uploading. We first randomly selected 75 positive and 75 negative images, then 60% from the total of 150 images used for training the SVM classifier and use the remaining 40% of images to test the performance of the trained model counting the number of success and failure, after that repeat the process 30 times, then calculate the average of the accuracy, sensitivity and specificity.

**Figure 2.**
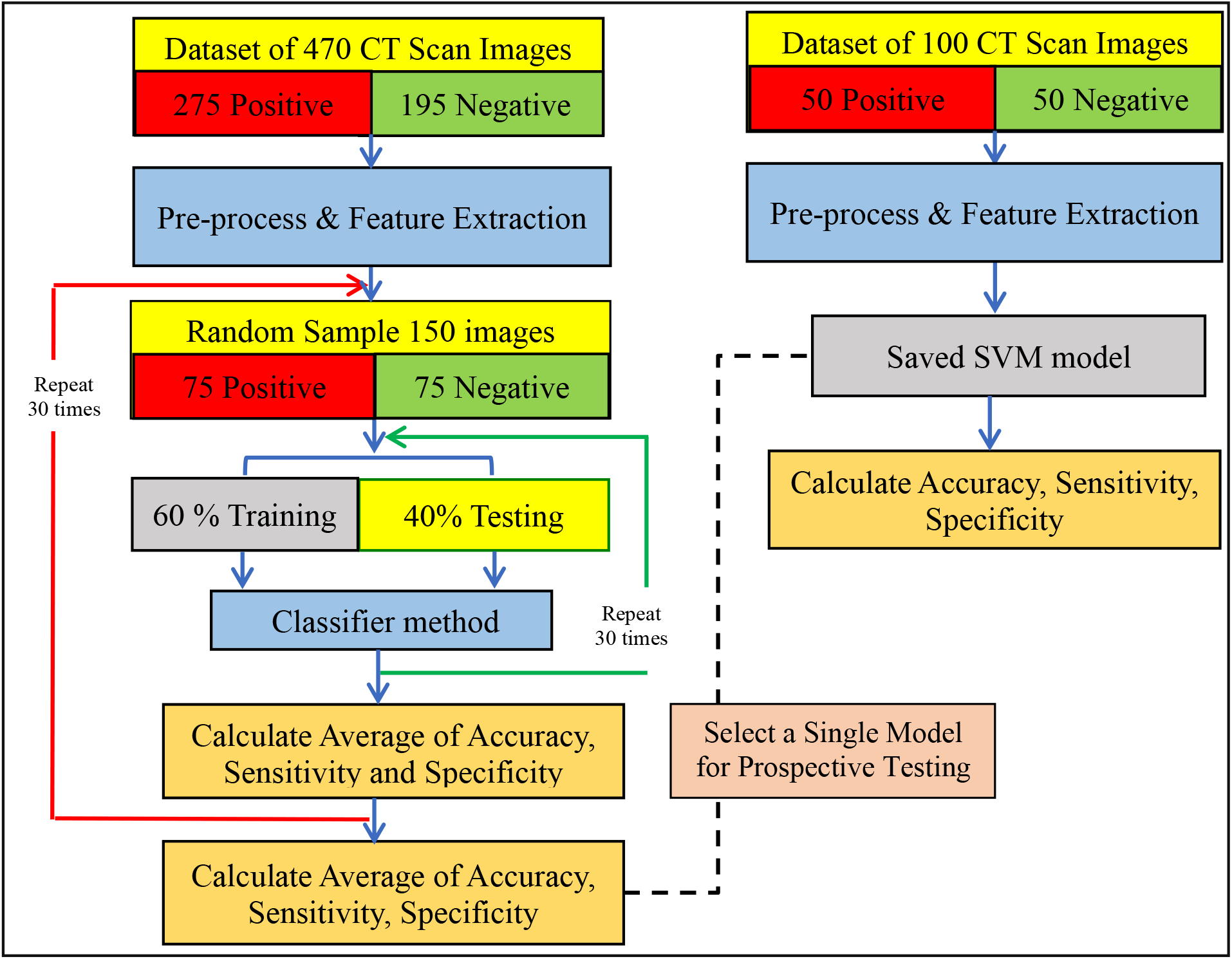
Experimental design and protocol.

In order to reduce the chance of biasness we repeat the experiment 30 times then find the average of that to produce the accuracy, sensitivity and specificity as shown in Table 1, below. To justify and explain our choice of the FFT-Gabor feature representation of CT scan images,

**Table 1.**
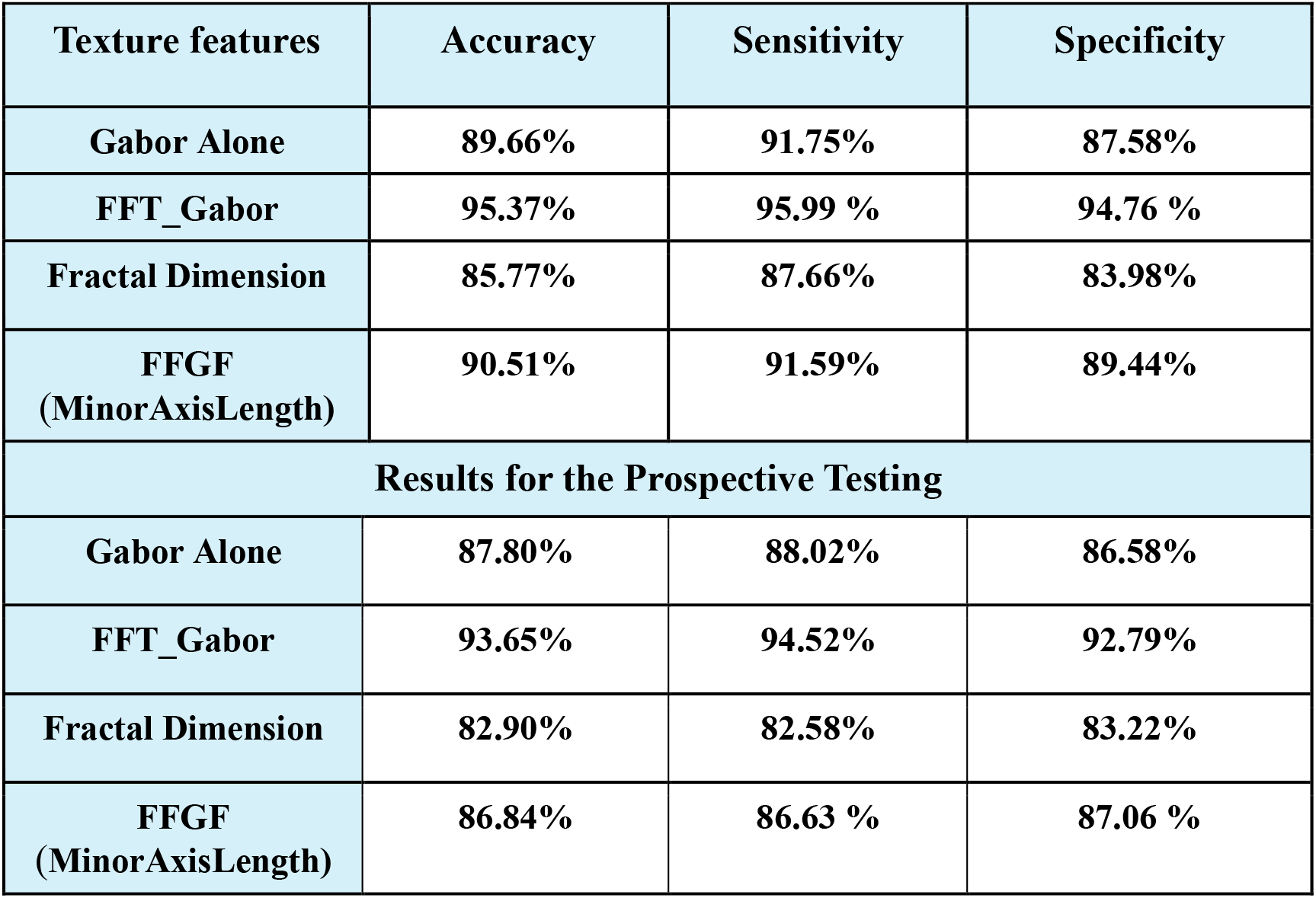
Classification result based on different texture features using SVM classifier.

we repeated the same experiments with other known texture features including but not limited to: Fractal dimension [13], Gabor filter in the spatial domain, the Fast Fourier Geometric Features (but only using the length of the Minor Axis of the best binary ellipse fitted to the spectrum image post adaptive thresholding) [13]. The performances of few other tested texture features are comparable to, or slightly lower than, the lowest performing schemes in this list.

The results not only confirm superiority of the FFT-Gabor scheme among other texture features, it also compares very well with the results, obtained manually by highly experienced clinicians, published by Ai, et al [5]. These results compare well with the accuracy achieved by the deep learning scheme of [6]. Indeed all the schemes outperform the one in [6] when tested on the original 470 images, with the FFT-Gabor scheme resulting in 10% absolute increased accuracy. For the prospective test, all the schemes show no need to retrain the system at this stage. Although, the Deep learning scheme developed in [9] is designed for slightly different purpose of characterising COVID-19 pneumonia our schemes also outperform that model. To have a more credible comparison with the work in [9] we must use their dataset.

Despite the obvious potential of chest CT-Scan, leading radiologists and clinicians expressed concern about the risk of viral transmission during patient transport to the imaging department posing an added risk to the medical staff and other patients. Moreover, the extensive efforts needed to disinfect the instruments between patients have a slow-down effect on the process and bring to question its advantage as an efficient tool in the fight against the Virus. However, CT Scan may play a critical role in the diagnosis of asymptomatic and pre-symptomatic cases where PCR test is not reliable or conclusive enough. Hence, it is essential for clinicians to have as many tools as possible in order to differentiate between positive and negative COVID-19. Moreover, accurate identification is also important to determine the optimal care management for these patients.

## 4. Conclusion and Future work

We have provided evidences in support of using automatic machine learning for texture analysis of Chest CT scans of suspected COVID-19 patients to complement existing RT-PCR lab testing. This could provide a reliable method of diagnosis in critical cases where the PCR test is not conclusive enough especially in pre-symptomatic patients. Since more samples are frequently uploaded, then we shall continue to upgrade our results by conducting more prospective experiments with new samples. It is also our intention is to conduct similar investigations if and when ultrasound lung scans become available in the future.

## Data Availability

non

https://github.com/UCSD-AI4H/COVID-CT

